# Diabetic Retinopathy: Knowledge, Staging and Barriers to Early Presentation in a Tertiary Eye Care Centre in Ghana

**DOI:** 10.1101/2024.12.01.24318265

**Authors:** Emmanuel Appiagyei, Yaw Akye Essuman, Akwasi Agyeman Ahmed

**Affiliations:** Eye Department, Komfo Anokye Teaching Hospital, Kumasi, Ghana; Department of Optometry and Visual Science, Kwame Nkrumah University of Science and Technology, Kumasi, Ghana; Lions International Eye Centre, Korle Bu Teaching Hospital, Accra, Ghana

**Keywords:** Diabetes mellitus, Diabetic retinopathy, ETDRS classification, Visual impairment, Ghana

## Abstract

**Background:** Diabetic retinopathy is the most common microvascular complication of diabetes mellitus and a leading cause of new-onset blindness in populations of working age. Late presentation of patients to eye care facilities has been associated with the development of vision-threatening complications. The study sought to determine the clinical profile, knowledge and factors influencing late presentation of diabetic retinopathy among diabetics at the Eye Centre of the Komfo Anokye Teaching Hospital in Ghana.

**Methods:** This descriptive cross-sectional study was conducted between the periods of February 2022 to April 2022. All diabetic patients referred to the retina clinic were examined for diabetic retinopathy. Both eyes of participants were examined and the eye with the most severe form of diabetic retinopathy was used for the staging. A structured questionnaire was employed to collect patients’ socio-economic factors and knowledge on diabetic retinopathy. Data was analysed using the SPSS ver. 23 software. Statistical significance was set at p<0.05.

**Results:** A total of 78 diabetic patients were included in the study. Thirty-six (36, 46.2%) were males and 42 (53.8%) females. Mean age of the study population was 49.9±11.4 years. The mean (+SD) knowledge score of study participants on diabetic retinopathy was 5.3 (±3.2) with a maximum possible score of 12. Majority of the participants (65%) were aware that uncontrolled diabetes mellitus affects the eyes; 7.7% had mild NPDR, 16.7%) had moderate NPDR, 10.3%; severe NPDR, 19.2%; very severe NPDR, 9.0%; high risk PDR and 37.2% showed signs of advanced PDR. Fifty-four participants (69.2%) highlighted a lack of knowledge about the condition as the main reason for late presentation.

**Conclusion:** Most diabetic patients who presented to the eye clinic had advanced PDR stage of the condition. The major factor to late presentation of diabetic retinopathy cases was lack of knowledge about the condition.

## Introduction

Diabetic retinopathy (DR) is a major complication of diabetes mellitus (DM), which remains a leading cause of visual loss in working-age populations in developing countries [1,2]. The diagnosis of DR is made by clinical manifestations of vascular abnormalities in the retina. The Early Treatment Diabetic Retinopathy Study (ETDRS) classification is considered the gold standard in grading diabetic retinopathy [3]. Two main stages: the non-proliferative diabetic retinopathy (NPDR) and the proliferative diabetic retinopathy (PDR) stages, have been considered, with differences based on the presence of microaneurysms, intraretinal haemorrhages and neovascularization [4]. The latter stage may be associated with severe visual impairment due to development of complications such as vitreous haemorrhage, tractional retinal detachment and maculopathy [1,5].

Loss of vision is a long-term complications of diabetes mellitus. Globally, the total number of people with diabetes mellitus is estimated to rise from 366 million in 2011 to 552 million by 2030 with nearly 80% of this population being in the middle age or working group [6]. DR is the most common complication in type I diabetes mellitus and nearly all patients will have some degree of retinopathy 15–20 years after diagnosis. Similarly, more than 60% of type II diabetes patients will have evidence of DR during this period [7]. A meta-analysis of the overall global prevalence of diabetic retinopathy (DR) was reported to be 34.6% [8]. Studies done in Ghana and Nigeria have reported the prevalence of diabetic retinopathy between the ranges of 15.5% to 17% [9,10].

Key risk factors leading to the development and progression of DR are long-term DM, inadequate glycaemic control, hypertension (HTN), dyslipidaemia, nephropathy and gender [7]. Control of the modifiable risk factors through periodic eye examinations and appropriate interventions has been shown to impede the advancement of DR [7].

Visual impairment as a result of DR has a significant impact on patients’ quality of life, and can compromise their ability to manage their disease successfully, which can in turn have a negative impact on the incidence of other diabetic complications and overall life expectancy [11]. Several studies have reported that most patients with diabetes mellitus are aware of the detrimental effect that the condition may have on their sight but yet they do not routinely seek eye care [12–14]. Reported barriers to screening and regular ocular examinations include: lack of knowledge about diabetic retinopathy and non-compliance with screening guidelines by ophthalmologists due to socioeconomic, cultural, and geographic reasons [15–17].

## Materials and methods

This hospital based cross-sectional study was conducted from 1^st^ February 2022 to 30^th^ April 2022 at the Eye Center, Komfo Anokye Teaching Hospital (KATH) – a major eye referral center in Ghana. Patients aged 18 years and above, with confirmed cases of diabetes mellitus, were considered eligible for this study whereas diabetic retinopathy patients with other retinal vascular pathologies such as: sickle cell retinopathy, hypertensive retinopathy, retinal vasculitis, were excluded. A purposive sampling method was employed to recruit 78 diabetic retinopathy patients (156 eyes) at the study site. All study participants had their visual acuity measured with the LogMAR Chart. Objective and subjective refraction was performed for all participants and best corrected visual acuity (BCVA) was recorded. A slit lamp biomicroscope (Inami & Co, Japan) was used to examine the anterior segment of the eye and pupillary reaction test. Intraocular pressure was measured using a Goldmann Applanation Tonometer (Haag-Streit AG, Koeniz, Switzerland). The pupils of all participants were dilated with 1% tropicamide and a detailed posterior segment examination as much as the media clarity allowed was performed by a retina specialist (AAA). A stereoscopic examination of the disc and macula with a slit lamp and 78D Volk lens (Volk Optical, USA); as well as indirect ophthalmoscopy with a 20D lens (Volk Optical, USA) was used for posterior segment examination; clinical staging of DR was done. Findings were recorded on the data collection sheet. Information on duration of diabetes mellitus diagnosis, and the presence of comorbidities such as hypertension was assessed from the history. This study adhered strictly to the principles of the Declaration of Helsinki and was approved by the Institutional Review Board of the Komfo Anokye Teaching Hospital (KATH IRB/AP/028/22). A written Informed consent was obtained from all participants.

## Data Analysis

Statistical analysis was performed using SPSS Version 23.0 (IBM Corp., Armonk, NY). Descriptive analysis was performed using frequencies, percentages, means and standard deviations. Participant’s knowledge of DR was assessed by 17 questions with a maximum score of 12 points. A numerical value of 1 for correct response and 0 for incorrect response was given. Participants who scored greater than or equal to the mean (≥5.32) of knowledge questions were considered to have good knowledge and those who scored below the mean were considered as having poor knowledge. The Chi-square test was used to determine statistical significance between categorical variables. Statistical significance was set at p<0.05.

## Results

A total of 78 patients were included in the study. The mean age of the study population was 49.9±11.4 years with the minimum age of 32 years and maximum age of 79 years. Almost all the study participants were enrolled on the National health insurance scheme (83.3%) and served as the only means of accessing healthcare. Almost 90% of the study population had at least primary education. Details pertaining to the socioeconomic characteristics of participants can be found in Table 1.

**Table 1:** Sociodemographic characteristics and mean knowledge score of study participants.

| Characteristics | Frequency<br>(%) | Mean | P-value |
| --- | --- | --- | --- |
|  |  | knowledge<br>score (SD) |  |
| Total | 78 (100.0) | 5.3 (3.2) |  |
| Age group, years |  |  | 0.212 |
| <40 | 19 (24.4) | 5.47 (3.66) |  |
| 40-49 | 19 (24.4) | 4.58 (2.89) |  |
| 50-59 | 30 (38.4) | 5.23 (3.10) |  |
| 60-69 | 5 (6.4) | 8.40 (1.95) |  |
| ≥70 | 5 (6.4) | 4.80 (3.35) |  |
| Gender |  |  | 0.506 |
| Male | 36 (46.2) | 5.58 (3.43) |  |
| Female | 42 (53.8) | 5.10 (3.01) |  |
| Occupation |  |  | 0.163 |
| Trader | 26 (33.3) | 4.81 (2.87) |  |
| Artisan | 13 (16.7) | 5.08 (3.30) |  |
| Farmer | 5 (6.4) | 4.60 (2.30) |  |
| Professional | 14 (17.9) | 7.43 (2.68) |  |
| Unemployed | 7 (9) | 4.29 (3.04) |  |
| Other | 13 (16.7) | 5.15 (4.10) |  |
| Marital status |  |  | 0.952 |
| Single | 11 (14.1) | 5.64 (3.72) |  |
| Married | 50 (64.1) | 5.16 (3.31) |  |
| Divorced | 7 (9) | 5.57 (3.51) |  |
| Widowed | 10 (12.8) | 5.60 (2.01) |  |
| Health insurance |  |  | 0.063 |
| NHIS | 65 (83.3) | 5.22 (3.11) |  |
| Private insurance schemes | 6 (7.7) | 8.00 (1.79) |  |
| None | 7 (9) | 4.00 (4.00) |  |
| Educational level |  |  | 0.594 |
| Primary | 10 (12.8) | 4.90 (3.21) |  |
| JHS | 23 (29.5) | 5.17 (3.28) |  |
| SHS | 12 (15.4) | 5.33 (3.08) |  |
| Tertiary | 25 (32.1) | 6.04 (3.49) |  |
| None | 8 (10.3) | 4.00 (2.14) |  |
| Housing type |  |  | 0.314 |
| Single room | 5 (6.4) | 4.00 (1.87) |  |
| Chamber and hall | 24 (30.8) | 4.63 (3.23) |  |
| Self-contained house | 38 (48.7) | 5.97 (3.05) |  |
| Apartment/Flat | 11 (14.1) | 5.18 (3.20) |  |

NHIS=National Health Insurance Scheme, JHS=Junior High School, SD=Standard Deviation, SHS=Senior High School

The mean (+SD) knowledge score of study participants about DR was 5.3 (±3.2) with a maximum possible score of 12. There was no statistically significant association between the mean scale scores of knowledge of diabetic retinopathy and sociodemographic characteristics of participants.

Participants’ level of knowledge in the various sections of diabetic retinopathy is represented in Table 2.

**Table 2:**
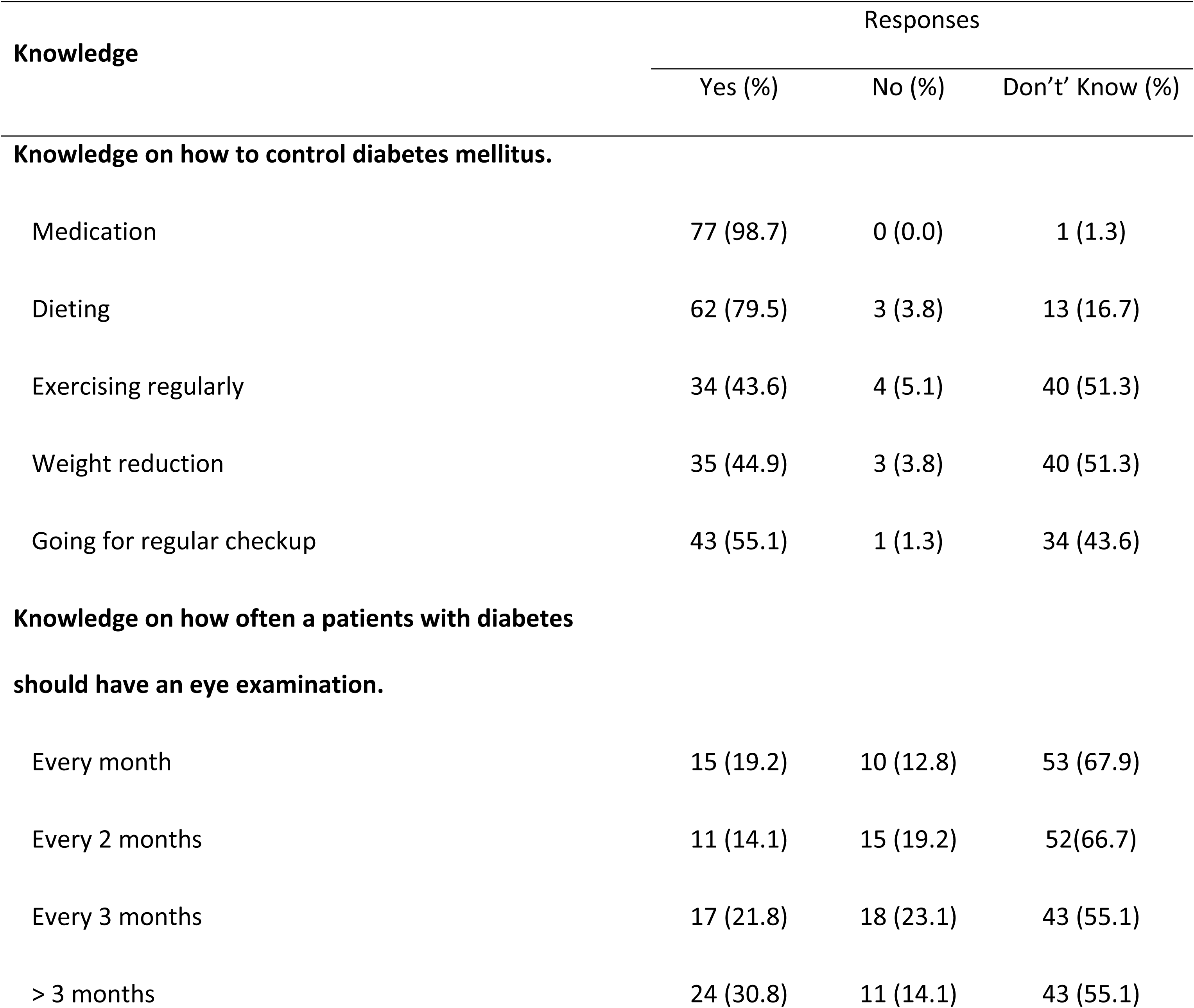
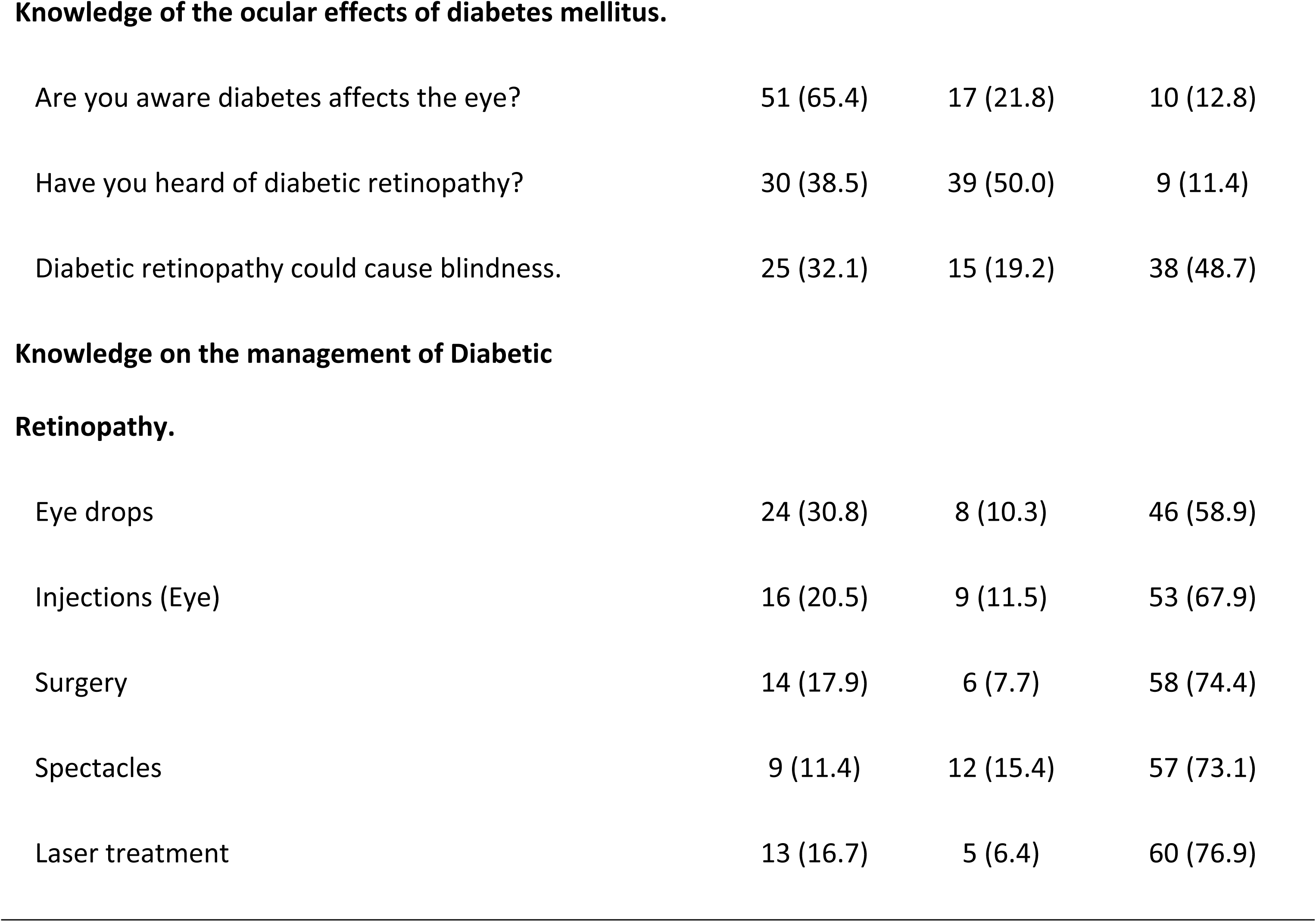
Knowledge of study participants on diabetic retinopathy (N=78).

Hundred and fifty six eyes (78 right eyes and 78 left eyes) were examined. When both eyes showed different stages of DR, a diagnosis was made based on the eye showing the most advanced stage of the condition. All participants (78, 100%) had at least one form of DR in one or both eyes. Among the 78 participants that were examined, 6 (7.7%) showed signs of mild NPDR, 13 (16.7%) showed signs of moderate NPDR, 8 (10.3%), severe NPDR; 15 (19.2%), very severe NPDR; 7 (9.0%), high risk PDR and 29 (37.2%) showed signs of advanced PDR. (Table 3).

**Table 3:**
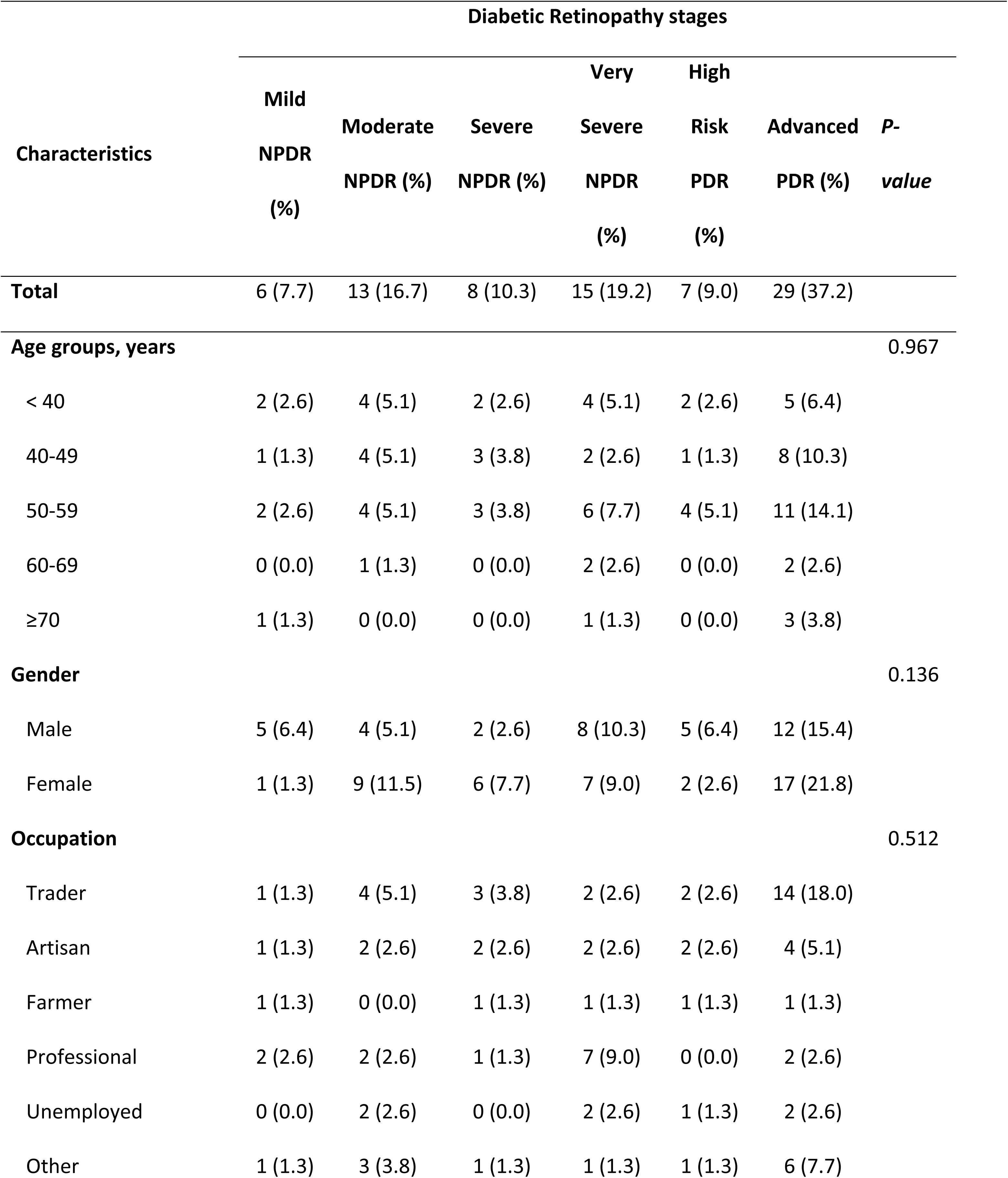
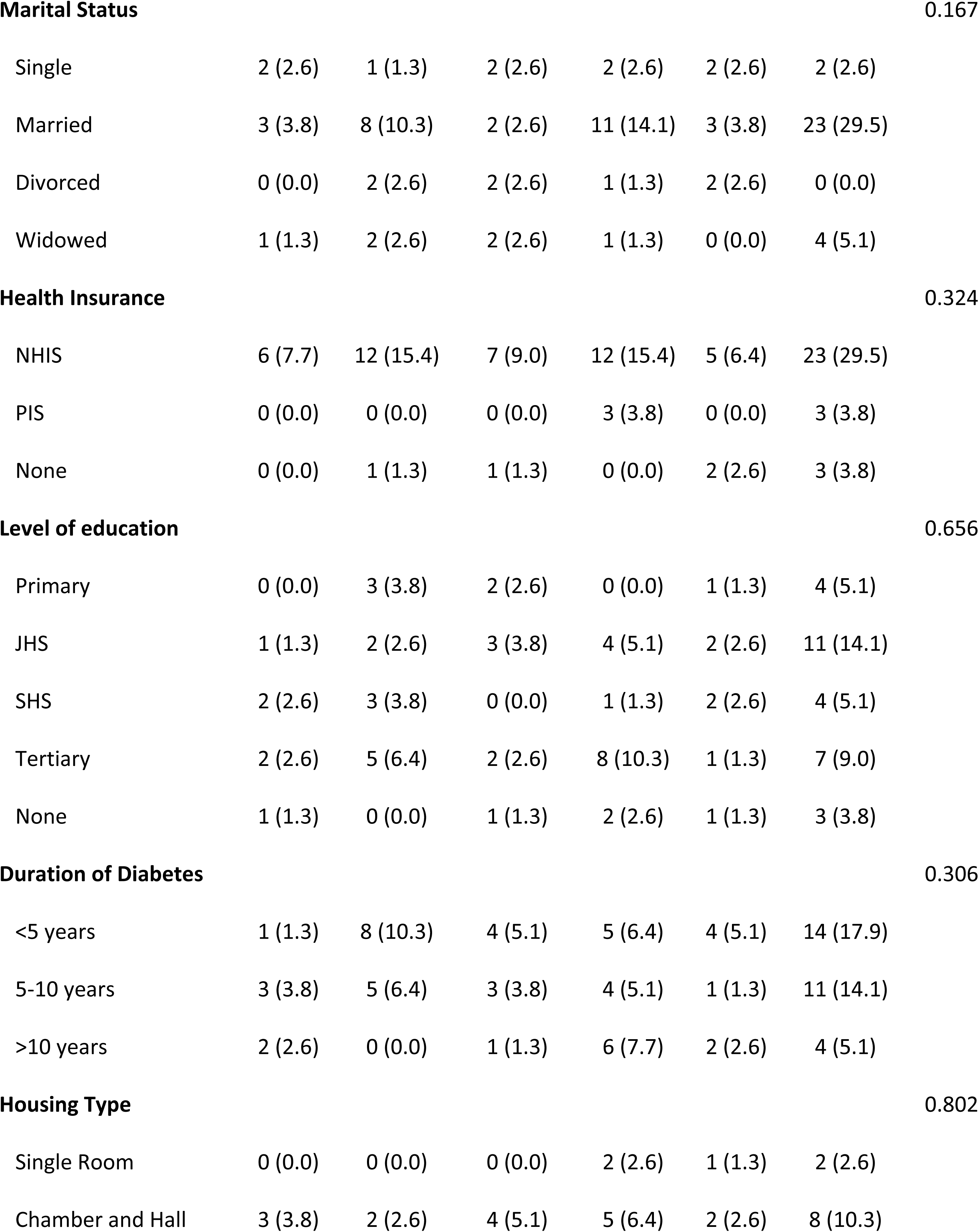
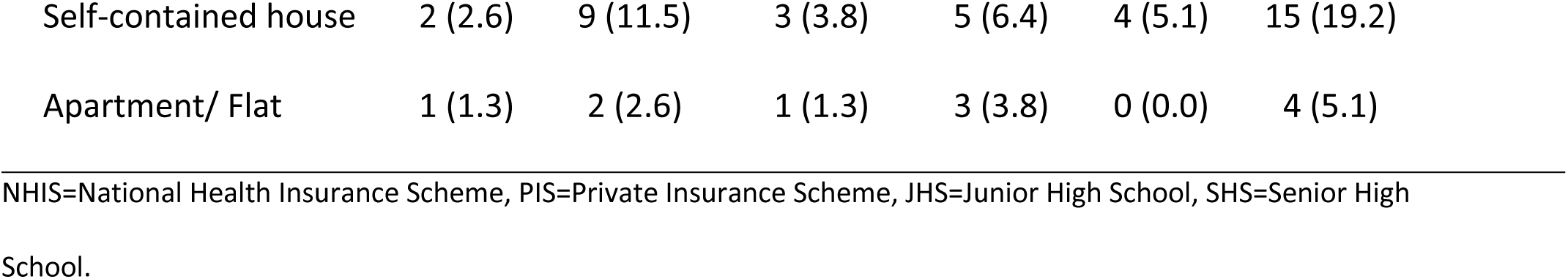
Stages of Diabetic Retinopathy among study participants.

| Characteristics | Diabetic Retinopathy stages |  |  |  |  |  | P-value |
| --- | --- | --- | --- | --- | --- | --- | --- |
|  | Mild<br>NPDR<br>(%) | Moderate<br>NPDR (%) | Severe<br>NPDR (%) | Very | High | Advanced<br>PDR (%) |  |
|  |  |  |  | Severe | Risk |  |  |
|  |  |  |  | NPDR<br>(%) | PDR<br>(%) |  |  |
| Total | 6 (7.7) | 13 (16.7) | 8 (10.3) | 15 (19.2) | 7 (9.0) | 29 (37.2) |  |
| Age groups, years |  |  |  |  |  |  | 0.967 |
| < 40 | 2 (2.6) | 4 (5.1) | 2 (2.6) | 4 (5.1) | 2 (2.6) | 5 (6.4) |  |
| 40-49 | 1 (1.3) | 4 (5.1) | 3 (3.8) | 2 (2.6) | 1 (1.3) | 8 (10.3) |  |
| 50-59 | 2 (2.6) | 4 (5.1) | 3 (3.8) | 6 (7.7) | 4 (5.1) | 11 (14.1) |  |
| 60-69 | 0 (0.0) | 1 (1.3) | 0 (0.0) | 2 (2.6) | 0 (0.0) | 2 (2.6) |  |
| ≥70 | 1 (1.3) | 0 (0.0) | 0 (0.0) | 1 (1.3) | 0 (0.0) | 3 (3.8) |  |
| Gender |  |  |  |  |  |  | 0.136 |
| Male | 5 (6.4) | 4 (5.1) | 2 (2.6) | 8 (10.3) | 5 (6.4) | 12 (15.4) |  |
| Female | 1 (1.3) | 9 (11.5) | 6 (7.7) | 7 (9.0) | 2 (2.6) | 17 (21.8) |  |
| Occupation |  |  |  |  |  |  | 0.512 |
| Trader | 1 (1.3) | 4 (5.1) | 3 (3.8) | 2 (2.6) | 2 (2.6) | 14 (18.0) |  |
| Artisan | 1 (1.3) | 2 (2.6) | 2 (2.6) | 2 (2.6) | 2 (2.6) | 4 (5.1) |  |
| Farmer | 1 (1.3) | 0 (0.0) | 1 (1.3) | 1 (1.3) | 1 (1.3) | 1 (1.3) |  |
| Professional | 2 (2.6) | 2 (2.6) | 1 (1.3) | 7 (9.0) | 0 (0.0) | 2 (2.6) |  |
| Unemployed | 0 (0.0) | 2 (2.6) | 0 (0.0) | 2 (2.6) | 1 (1.3) | 2 (2.6) |  |
| Other | 1 (1.3) | 3 (3.8) | 1 (1.3) | 1 (1.3) | 1 (1.3) | 6 (7.7) |  |
| **Marital Status** |  |  |  |  |  |  | 0.167 |

|  |  |  |  |  |  |  |
| --- | --- | --- | --- | --- | --- | --- |
| Single | 2 (2.6) | 1 (1.3) | 2 (2.6) | 2 (2.6) | 2 (2.6) | 2 (2.6) |
| Married | 3 (3.8) | 8 (10.3) | 2 (2.6) | 11 (14.1) | 3 (3.8) | 23 (29.5) |
| Divorced | 0 (0.0) | 2 (2.6) | 2 (2.6) | 1 (1.3) | 2 (2.6) | 0 (0.0) |
| Widowed | 1 (1.3) | 2 (2.6) | 2 (2.6) | 1 (1.3) | 0 (0.0) | 4 (5.1) |
| **Health Insurance** |  |  |  |  |  |  | 0.324 |

|  |  |  |  |  |  |  |
| --- | --- | --- | --- | --- | --- | --- |
| NHIS | 6 (7.7) | 12 (15.4) | 7 (9.0) | 12 (15.4) | 5 (6.4) | 23 (29.5) |
| PIS | 0 (0.0) | 0 (0.0) | 0 (0.0) | 3 (3.8) | 0 (0.0) | 3 (3.8) |
| None | 0 (0.0) | 1 (1.3) | 1 (1.3) | 0 (0.0) | 2 (2.6) | 3 (3.8) |
| **Level of education** |  |  |  |  |  |  | 0.656 |

|  |  |  |  |  |  |  |
| --- | --- | --- | --- | --- | --- | --- |
| Primary | 0 (0.0) | 3 (3.8) | 2 (2.6) | 0 (0.0) | 1 (1.3) | 4 (5.1) |
| JHS | 1 (1.3) | 2 (2.6) | 3 (3.8) | 4 (5.1) | 2 (2.6) | 11 (14.1) |
| SHS | 2 (2.6) | 3 (3.8) | 0 (0.0) | 1 (1.3) | 2 (2.6) | 4 (5.1) |
| Tertiary | 2 (2.6) | 5 (6.4) | 2 (2.6) | 8 (10.3) | 1 (1.3) | 7 (9.0) |
| None | 1 (1.3) | 0 (0.0) | 1 (1.3) | 2 (2.6) | 1 (1.3) | 3 (3.8) |
| **Duration of Diabetes** |  |  |  |  |  |  | 0.306 |

|  |  |  |  |  |  |  |
| --- | --- | --- | --- | --- | --- | --- |
| <5 years | 1 (1.3) | 8 (10.3) | 4 (5.1) | 5 (6.4) | 4 (5.1) | 14 (17.9) |
| 5-10 years | 3 (3.8) | 5 (6.4) | 3 (3.8) | 4 (5.1) | 1 (1.3) | 11 (14.1) |
| >10 years | 2 (2.6) | 0 (0.0) | 1 (1.3) | 6 (7.7) | 2 (2.6) | 4 (5.1) |
| **Housing Type** |  |  |  |  |  |  | 0.802 |

|  |  |  |  |  |  |  |
| --- | --- | --- | --- | --- | --- | --- |
| Single Room | 0 (0.0) | 0 (0.0) | 0 (0.0) | 2 (2.6) | 1 (1.3) | 2 (2.6) |
| Chamber and Hall | 3 (3.8) | 2 (2.6) | 4 (5.1) | 5 (6.4) | 2 (2.6) | 8 (10.3) |
| Self-contained house | 2 (2.6) | 9 (11.5) | 3 (3.8) | 5 (6.4) | 4 (5.1) | 15 (19.2) |
| Apartment/ Flat | 1 (1.3) | 2 (2.6) | 1 (1.3) | 3 (3.8) | 0 (0.0) | 4 (5.1) |

NHIS=National Health Insurance Scheme, PIS=Private Insurance Scheme, JHS=Junior High School, SHS=Senior High School.

When the eyes of participants were analyzed separately, it was found that, out of the 156 eyes, 138 (88.5%) had DR and 18 (11.5%) had no DR. In the analysis of only the right eye (OD) of participants, 65 (83.3%) had DR and 13 (16.7%) did not have DR, whereas in the left eye (OS) of study participants, 73 (93.6%) had DR and 5 (6.4%) did not have DR. (Figure 1).

**Figure 1:**
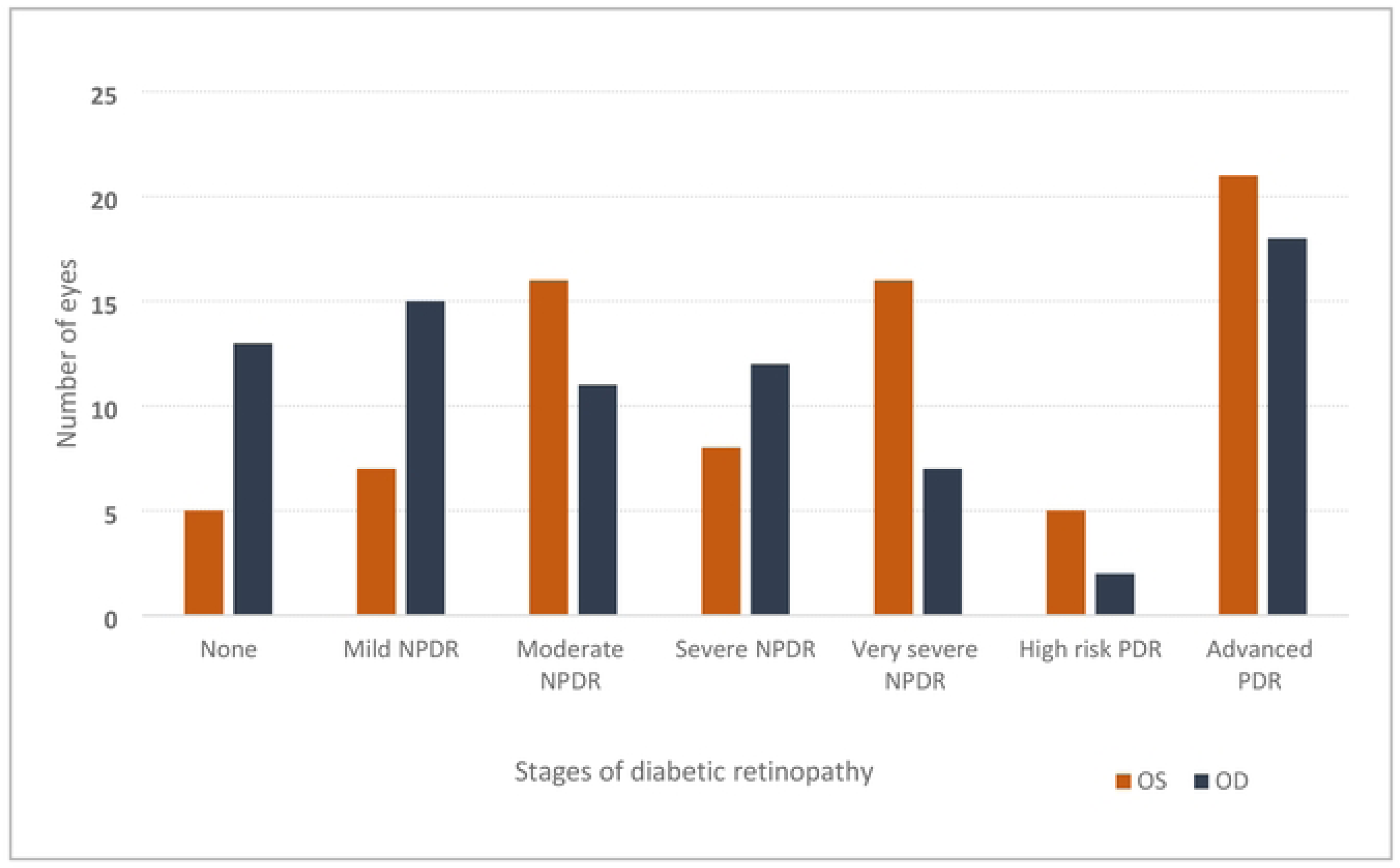
Stages of diabetic retinopathy in both eyes of study participants (N=78).

Factors elucidated for late presentation among study participants include: lack of knowledge about diabetic retinopathy (54, 69.2%), lack of access to eye care (15, 22.1%), cost/insurance (17, 21.8%), fear of discovery (25, 32.1%), transportation difficulties (3, 3.8%) and others (7, 9.0%).

## Discussion

The mean age of our study population; patients with diabetes (50 years) was similar to other hospital-based studies conducted by in Jordan [18] and Turkey [19], but unlike other developing countries [20,21], where patients were much younger. This may be due to late detection of diabetes or late presentation for eye examination among our study population.

More females than males were seen during the study period, this was similar to the study in Turkey which reported a preponderance of female participants (51.8%) among their study population [19]. This was contrary to findings seen in studies in Jordan where male prevalences of 50.4%, and 54.9%, were recorded [22]. The higher percentage of females found in this study could be attributed to the recent upward trends in the prevalence of diabetes among women [6].

Majority of the study population were traders (33.3%) followed by professionals (17.9%). Regarding level of education, majority of the study population had attained tertiary education, and most of the study population occupied either a chamber/hall (30.8%) or self-contained house (48.7%). These socio-demographic characteristics reflects on the fraction of population that have higher prevalence of diabetes as reported in prevalence studies (diabetes) in the country [9]. A study in an adult Ghanaian diabetic population found that diabetes is prevalent among adults aged 50 years and above, females, higher level of education, wealth households and professionals [23]. These findings reflect in the sociodemographic characteristics of our study population in this study.

On assessment of the knowledge of participants on diabetic retinopathy, we found that 65.4% of participants were aware that diabetes had ocular effects. Only 38.5% of them had heard about diabetic retinopathy and 32.1% knew DR could result in blindness. Contrary to our findings, another study found that 98.3% of the study participants knew that diabetes could affect eyes with 99.1% believing that it could lead to blindness, while 50.4% of them were aware of diabetic retinopathy [22]. A study carried out in Jordanese population also revealed that 88.2% of study participants were aware of the effects of diabetes on the eye with 81.9% stating that diabetes could lead to blindness [18]. These findings were also seen in Turkey, where a study also reported higher levels of knowledge among participants where 88.1% of the study participants being aware of the effects of diabetes on the eyes [19]. The disparities in the level of knowledge as reported in this study and other studies could be attributed to a lower educational level among our study populations. Also, there is an established national eye health care program and referral guidelines regarding DM and DR management for primary care in Jordan which positively influence knowledge about the conditions. Patients are referred for an eye examination at the same time as their diagnosis of DM. This may explain the high level of awareness of DR found in their study. The lower levels of knowledge regarding the awareness of diabetic retinopathy accounts for the lower mean scale score of (5.32) that was found in the study.

The stages of diabetic retinopathy were graded according to the ETDRS classifications. In the current study, 37.2% of the study population showed signs of advanced PDR, while 9.0% had high risk PDR and 19.2% had very severe NPDR. Several findings have been reported in other studies, including in Blantyre, Malawi, where 19.7% of type 2 DM cases were found to have STDR which was defined as pre-proliferative, proliferative and or the presence of macular edema, while a further 8.5% had PDR. They also found an STDR prevalence of 18.8% and PDR prevalence of 12.5% in type 1 diabetic participants [24]. A study in a Chinese population [25], found an STDR prevalence of 12.6% among 16305 participants, while another study [26] in Hangzhou, China, found that 80% of participants had STDR, including 67% of first-time attendees at the study site. Even though the method of classification in the other studies were different from this study, it was observed that the percentage of the study population at the stages of advanced PDR and high risk PDR was high (46.2%) as compared to the studies in Malawi and China [24,25]. The higher percentages recorded in our study could be attributed to the late presentation of diabetic patients to the eye clinic for examination, resulting in more of them presenting with advanced PDR stages.

Study subjects reported several factors that led to the late presentation of diabetic retinopathy cases. Lack of knowledge about diabetic retinopathy was the most reported factor (69.2%) followed by fear of discovery (32.1%), lack of access (22.1%), high costs (21.8) and transportation difficulties (3.8). These findings were similar to those seen in Owo, Nigeria, where lack of knowledge about DR was cited by 62.8% of participants [27]. Davalgi et al. (2018) also reported lack of knowledge about DR as the number one barrier to timely eye examination by diabetic patients [28]. Another study reported that cost of service delivery also hindered timely eye examination by diabetic patients [29]. Lack of access to eye examination as reported by some study participants may be due to an inadequate number of ophthalmologists in the country and the unequal distribution of eye care services in the country [30]. Most eye care centers are in the urban regions of the country which deprives diabetic patients in the remote areas from seeking eye care and even those that decide to make efforts to seek eye care are faced with transport difficulties.

This study had some limitations. It was a hospital-based study with a small sample size as compared to other studies and as such findings of the study may not be an exact representation of diabetic patients and diabetic retinopathy in the country. The study however, provides much-needed insight into factors influencing late presentation among diabetic retinopathy patients and should thus, serve as a foundation for future research and public health interventions on the subject.

## Conclusions

The study identified majority of diabetic patients having poor knowledge about diabetic retinopathy. Most diabetic patients were found to present to the eye clinic advanced PDR stages with many citing a lack of knowledge about the condition as the major factor for their late presentation. There is the need for compulsory ocular examination for all diabetic patients at their hospitals as well as wide scale education on diabetes mellitus and its ocular complications in remote areas.

## Data Availability

All relevant data are within the manuscript and its Supporting Information files

## List of abbreviations

DM: Diabetes mellitus
DR: Diabetic retinopathy
ETDRS: Early Treatment Diabetic Retinopathy study
KATH: Komfo Anokye Teaching Hospital
NPDR: Non proliferative diabetic retinopathy
OD: Oculus dexter
OS: Oculus sinister
PDR: Proliferative diabetic retinopathy
STDR: Sight threatening diabetic retinopathy

## Declarations

### Ethics approval and consent to participate

This study adhered strictly to the principles of the Declaration of Helsinki and was approved by the Institutional Review Board of the Komfo Anokye Teaching Hospital (KATH IRB/AP/028/22). A written Informed consent was obtained from all participants.

### Consent for publication

Not applicable.

### Availability of data and materials

All relevant data are within the manuscript and its Supporting Information files.

### Authors’ contributions

All authors contributed to the conceptualization and design of the study, collection and management of data, analysis and interpretation and drafting and revision of manuscript.

